# *M. tuberculosis* transmission dynamics in congregate settings: a genomic epidemiology study

**DOI:** 10.1101/2024.12.18.24318332

**Authors:** Katharine S. Walter, Everton Ferreira Lemos, Ana Paula Cavalcante Aires Alves, Gabriela Felix Chaves Ferreira, Vanessa Maruyama Martins Coutinho, Barun Mathema, Joshua L. Warren, Caroline Colijn, Ted Cohen, Julio Croda, Jason R. Andrews

## Abstract

**Background:** One barrier to intervening in the global tuberculosis (TB) pandemic is that it is unknown whether *M. tuberculosis* transmission largely occurs through repeated close exposures among few contacts or many shorter-term contacts. Identifying sources of transmission is particularly urgent in congregate settings with high incidence of infection.

*Methods:* To identify drivers of *M. tuberculosis* transmission within a congregate setting with extremely high incidence of TB, we conducted genomic surveillance in a prison system in Central West Brazil. We whole genome sequenced *M. tuberculosis* isolates and collected detailed incarceration histories, including movements between and within prisons. We integrated incarceration histories with *M. tuberculosis* genomes to investigate the relationship between exposures of differing proximity (cell, cell block, prison) and transmission risk, using genomic clustering as a proxy for transmission.

*Findings:* We collected detailed incarceration histories for 595 individuals from whom we sequenced 561 high quality *M. tuberculosis* genomes. A month-long increase in exposure to an individual with TB within a prison cell increased the odds of pairwise genomic clustering by 7.4% (95% CI: 4.4-10.4%) and a six-month increase in exposure, by 54% (95% CI: 29.9%-82.5%). Most (89%; 528 of 595) individuals with TB had at least one potential week-long exposure in a prison cell to another individual with TB, and frequently many, with a median of 12 (IQR: 5-21) potential unique exposures to individuals in prison cells. Frequent movements by the prison system create a highly connected contact network: individuals with TB were transferred a median of 5 (IQR: 1-17) times in the 12 months before diagnosis.

*Interpretation:* While close exposures within a prison were related to pairwise genomic clustering, most individuals with TB had multiple exposures to other individuals with TB due to frequent movements by the prison system. Our results support the urgent expansion of prison-wide mass screenings, TB preventive therapy, and structural interventions to reduce transmission risk in prisons and other congregate settings.

*Funding:* National Institutes of Health (NIAID: 5K01AI173385, R01AI100358, and R01AI149620)

**Research in context:** *Evidence before this study:* We searched PubMed for relevant articles published in English from database inception to November 26, 2024, using the terms “*Mycobacterium tuberculosis*”, “transmission,” “genom*,” and “congregate setting” or “prison.” We found several genomic epidemiology articles describing close genetic relatedness of *M. tuberculosis* sampled from prisons and the community. These earlier genomic epidemiology studies did not include individual-level exposure or movement information. We additionally identified two studies that conducted environmental sampling in congregate settings: one that identified *M. tuberculosis* DNA in bioaerosols in a primary care clinic and one from environmental swabs collected in a prison. Previous studies did not investigate the types of contacts driving transmission in high-incidence congregate settings.

*Added value of this study:* We conducted a genomic epidemiology study of *M. tuberculosis* transmission in a congregate setting with extremely high incidence of tuberculosis (TB): a state prison system in Central West, Brazil. We integrated *M. tuberculosis* genomes with detailed individual movement data to reconstruct transmission linkages and infer the types of contacts associated with transmission in a congregate setting. We found that close contacts within a prison—incarceration within the same prison cell—increase the likelihood of transmission. Further, the frequent movement of individuals within and between prisons creates large, highly connected large contact networks. The result is that individuals have many close contacts with other individuals with tuberculosis, such that any single potential exposure may not result in genetically linked cases.

*Implications of all the available evidence:* Together, our results suggest that close exposures to other individuals with TB increase transmission risk in congregate settings with high incidence of TB. Due to frequent transfers within prison systems, people may have close exposures to many individuals with TB, with the result that contact tracing investigations may not be effective in such settings. Our results support the urgent expansion of mass screenings, TB preventive therapy, and structural interventions to reduce transmission risk in prisons and other congregate settings.

## Introduction

In 2024, tuberculosis (TB) regained its status as the infectious disease responsible for the largest global mortality burden^1^. Reducing the global burden of TB urgently requires minimizing the number of incident *M. tuberculosis* infections. Yet unlike many respiratory pathogens, the long and variable latency period of *M. tuberculosis* infection makes it challenging to identify sources of transmission and thus intervene^2^. One barrier is that it is unknown whether *M. tuberculosis* transmission occurs through close contacts or risk is more diffuse across contacts. While historically, it was thought that most transmission occurs among highly exposed contacts, more recent work has indicated that transmission frequently occurs outside of households, in the community ^2–7^.

There is an urgent need to identify sources of transmission both in the general population and in congregate settings such as hospitals, prisons, and mines, where transmission risk is frequently elevated compared to surrounding communities^8^. Prisons and other detention centers put the estimated 11.5 million people currently incarcerated around the world at extremely high risk of TB ^9,10^. Globally, TB incidence in prisons is 10 times that outside prisons^10^. In the Americas, incarceration rates have dramatically risen in the past thirty years, and 2.4 million people are currently incarcerated. Here, the incidence of TB within prisons is 26.9 times that outside prisons^11^. This extreme risk reflects the severe overcrowding, unhygienic living conditions, prolonged confinement in cells with poor ventilation and sunlight, poor nutritional provisions, and limited access to primary healthcare TB diagnostics and nutrition within prisons^12,13^.

A key unanswered question in congregate settings, including prisons, is whether transmission risk occurs largely between close contacts or is diffuse and frequently occurs between more distant, often-termed casual, contacts. If transmission is driven by identifiable close contacts, this could support the use of contact tracing within prisons. TB testing might, for example, be prioritized for close contacts. Alternatively, if transmission risk frequently occurs among more distant contacts, or if transmission linkages cannot be determined due to a highly connected network, control might focus on agnostic, mass screening approaches and tuberculosis preventive treatment (TPT) for the entire prison population.^14^ Resolving the contribution of close and distant contacts to transmission could additionally support the use of structural interventions, such as increased ventilation, air filtration or UV radiation^12,15^.

Genomic epidemiology has been powerfully applied to reconstruct *M. tuberculosis* transmission linkages including the directionality of transmission across both low and high-incidence settings^2^. Similar approaches have been applied to bacterial, viral, and fungal pathogens to infer transmission networks within congregate settings such as hospitals, schools, and workplaces (e.g., ^16^). These previous studies have implicated specific locations within buildings, occupations, and activities associated with transmission which can be prioritized for infection control.

To identify the type of contacts associated with *M. tuberculosis* transmission risk within prisons, we conducted a genomic surveillance study of *M. tuberculosis* in a prison system in Central West Brazil with an extremely high incidence of TB. We integrated detailed incarceration histories with *M. tuberculosis* genomes to infer transmission linkages and quantify the contribution of close and distant contacts to transmission.

## Methods

### Epidemiological setting

Brazil’s incarceration rate has increased from 132 per 100,000 in 2000 to 381 per 100,000 in 2020 and its prison population has more than tripled, increasing from 232,755 to 811,707 over the same period^9^. In 2024, Brazil’s incarcerated population is the third highest in the world^9^. Brazil’s criminal law defines different regimes or stages of incarceration: closed regimes, in which people are incarcerated full-time; semi-open regimes, in which people work outside prisons and return at night; and open regimes, under which people serve sentences outside of prison, but are required to make periodic court appearances^17^.

The state of Mato Grosso do Sul, which has one of the highest incarceration rates in Brazil (771 per 100,000 individuals), has 28 closed prisons, 7 semi-open prisons, and 2 open prisons (Fig. 1). As of 2024, there were a total of 21,483 incarcerated individuals and a system occupancy of 12,885, an occupancy rate of 167%. The most common criminal charge is drug trafficking, which is listed as the crime for 37% of individuals in the state. The burden of TB in this prison system is one of the highest reported in the world: a recent mass screening study in three of the largest state prisons found a TB prevalence of 4,034 per 100,000 people^18^.

**Fig 1.**
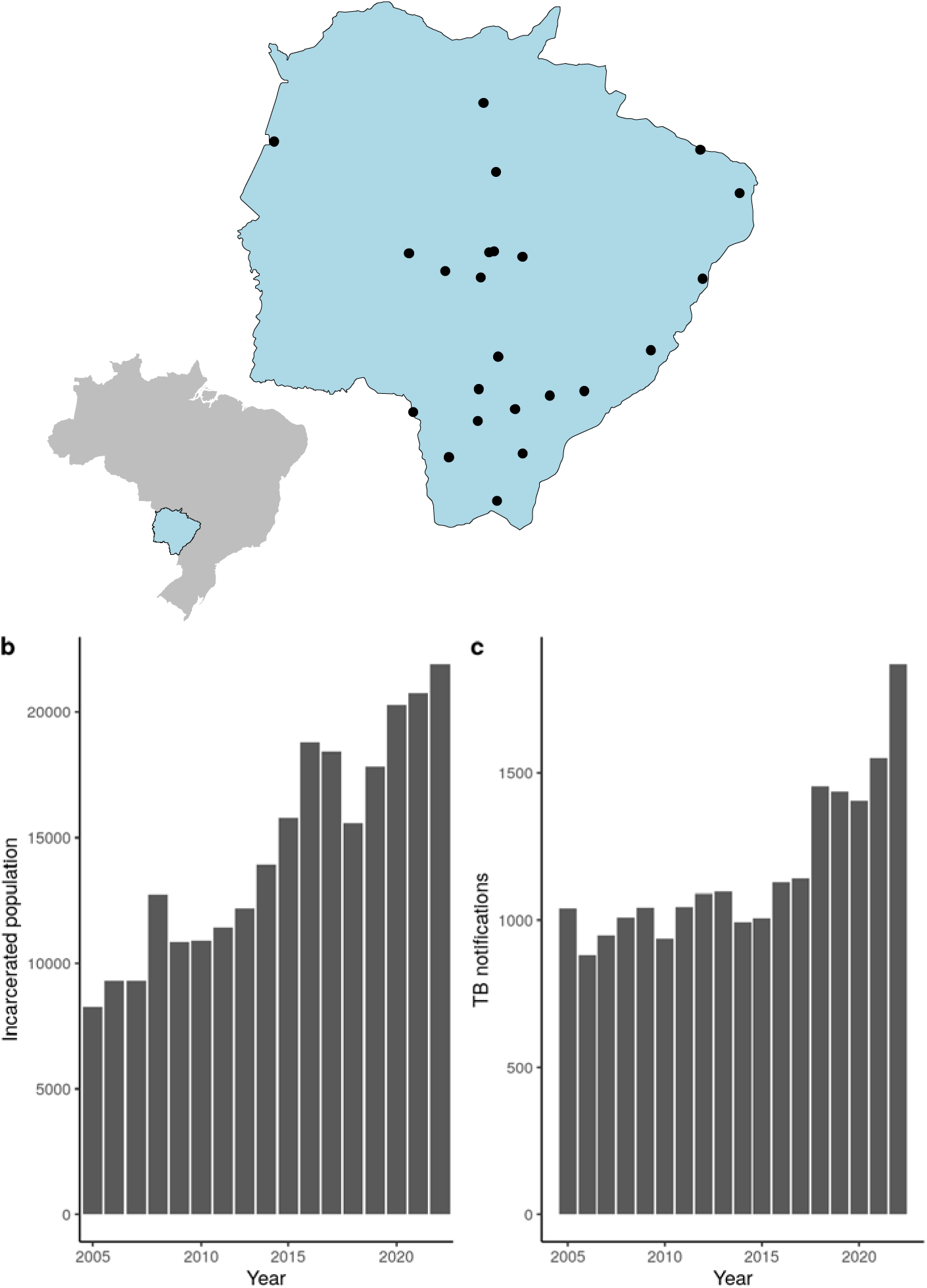
Study location. a) Map of Mato Grosso do Sul, with the location of the state’s 32 closed prisons shown as points. Inset is a map showing the location of Mato Grosso do Sul within Brazil. b) Total incarcerated population and c) number of TB notifications in Mato Grosso do Sul reported to Brazil’s national notifiable disease registry.

Prisons are often structured by blocks of cells surrounding open pavilions. Cells have narrow windows looking out to shared hallways and may have small, barred windows to the outside. Incarcerated individuals spend an average of 20 hours per day in cells and take meals within cells. During outdoor time, individuals are allowed to go into outdoor pavilions surrounded by blocks of cells where they are allowed to play sports or congregate.

### Genomic surveillance

We conducted population-based tuberculosis surveillance in the state of Mato Grosso do Sul in Central West Brazil from 2013 through 2022. Surveillance included active screening in three of the largest prisons in the state as well as ongoing passive surveillance by the state public health agency, LACEN. We sequenced whole genomes from cultured isolates on an Illumina NextSeq (2 x 151-bp). Sequence data for samples meeting quality filters are available on the Sequence Read Archive (SRA), in BioProject PRJNA671770.

### Variant identification and genomic clustering

We identified *M. tuberculosis* genomic variation from whole genome sequence data with a pipeline available at https://github.com/ksw9/mtb-call2^19^ (Supplementary Methods). Genomic clustering is often used as a proxy measure of recent *M. tuberculosis* transmission; isolates that are more closely genetically related are hypothesized to be more likely linked through recent transmission rather than travel-associated importation or re-activation of genetically distinct latent infections^20^. We applied a commonly used genetic distance thresholds of 5 or 12 SNPs to identify genomic clusters^20,21^.

### Incarceration history

To investigate shared exposures for incarcerated individuals with a TB notification, we obtained permission to access the Mato Grosso do Sul state penitentiary system administrative database (SIAPEN), which includes records for all individuals who have been contacted by the criminal justice system since 2013. Data are only available through manual query; we therefore searched SIAPEN with participant names and extracted information on sentencing and date and location of incarceration, including movements between prisons and movements within prisons. In some cases, movement data were incomplete. We truncated these final incarceration intervals at the median duration of incarceration between movements.

### Statistical analysis

We defined pairwise potential *M. tuberculosis* exposures as overlapping incarceration within a prison cell, prison block (group of cells surrounding a shared outdoor space), or prison. We defined potential exposures as the duration in days of overlapping incarceration with another individual with notified TB. We restricted exposures to those prior to the recipient’s date of TB notification. First, we used a logistic regression model to test whether an individual’s potential exposure in a prison cell, block, or prison was associated with an individual’s membership in a genomic cluster.

Second, we tested whether the pairwise duration of potential exposures within cells, blocks, and prisons were associated with shared *M. tuberculosis* genomic cluster with multivariable logistic regression. Cell-level exposures implied block- and prison-level exposures and block-level exposures implied prison-level exposures, therefore, we did not include explicit interaction terms. Paired or dyadic data, such as the pairwise potential exposures and pairwise *M. tuberculosis* genetic relatedness we analyze here, are not independent because individual members of pairs are present in multiple dyadic outcomes. To control for the non-independence of dyadic data, we fit a hierarchical Bayesian spatial modeling approach for dyadic pathogen genetic relatedness data with the R package *GenePair*^22^. We collected 50,000 samples from the joint posterior distribution after removing the first 10,000 samples for burn-in and thinning the remaining 40,000 samples by a factor of 10 to reduce posterior autocorrelation. We assessed convergence with Geweke’s diagnostic and present posterior means and 95% credible intervals (CIs)^22^ (Supplementary Methods).

### Ethical Approval

All participants provided written consent, and this study was conducted with the approval of the Research Ethics Committee from the Federal University of Grande Dourados, Federal University of Mato Grosso do Sul and National Research Ethics Committee (CONEP) (CAAE 37237814.4.0000.5160, 2676613.3.1001.5160, and 26620619.6.0000.0021) and Stanford University Institutional Review Board (IRB-40285) and the University of Utah (IRB_00177937).

## Results

### Within-prison and between-prison transfers are frequent

We identified 638 individuals who were diagnosed with TB during incarceration through active screening conducted by our group in three of the largest prisons in the state^18,24^ or through the state’s public health laboratory from 2013 to 2022, with an additional 34 records from 2007-2012. We manually queried the state incarceration database to assemble individual incarceration records that included date of incarceration and date and location of every movement within the state’s penitentiary system, including all carceral regimes and jails, and every exit from the system. We located incarceration records for 595 individuals and extracted a total of 21,030 individual movements.

We hypothesized that the criminal justice system’s frequent transfer of people within prisons and between prisons and jails could disseminate TB infection risk within institutions and across the state. Individuals were transferred a median of 30 times (interquartile range: 15-47), including movements within a prison, between prisons, or prison entries or exits. The median duration of incarceration between movements was 19 days (IQR: 6-65 days) after 2013, when cell-level information was more consistently recorded (Fig. 2). Individuals were moved a median of 5 times (IQR: 1-17) in the 12 months before their diagnosis with TB.

**Fig. 2.**
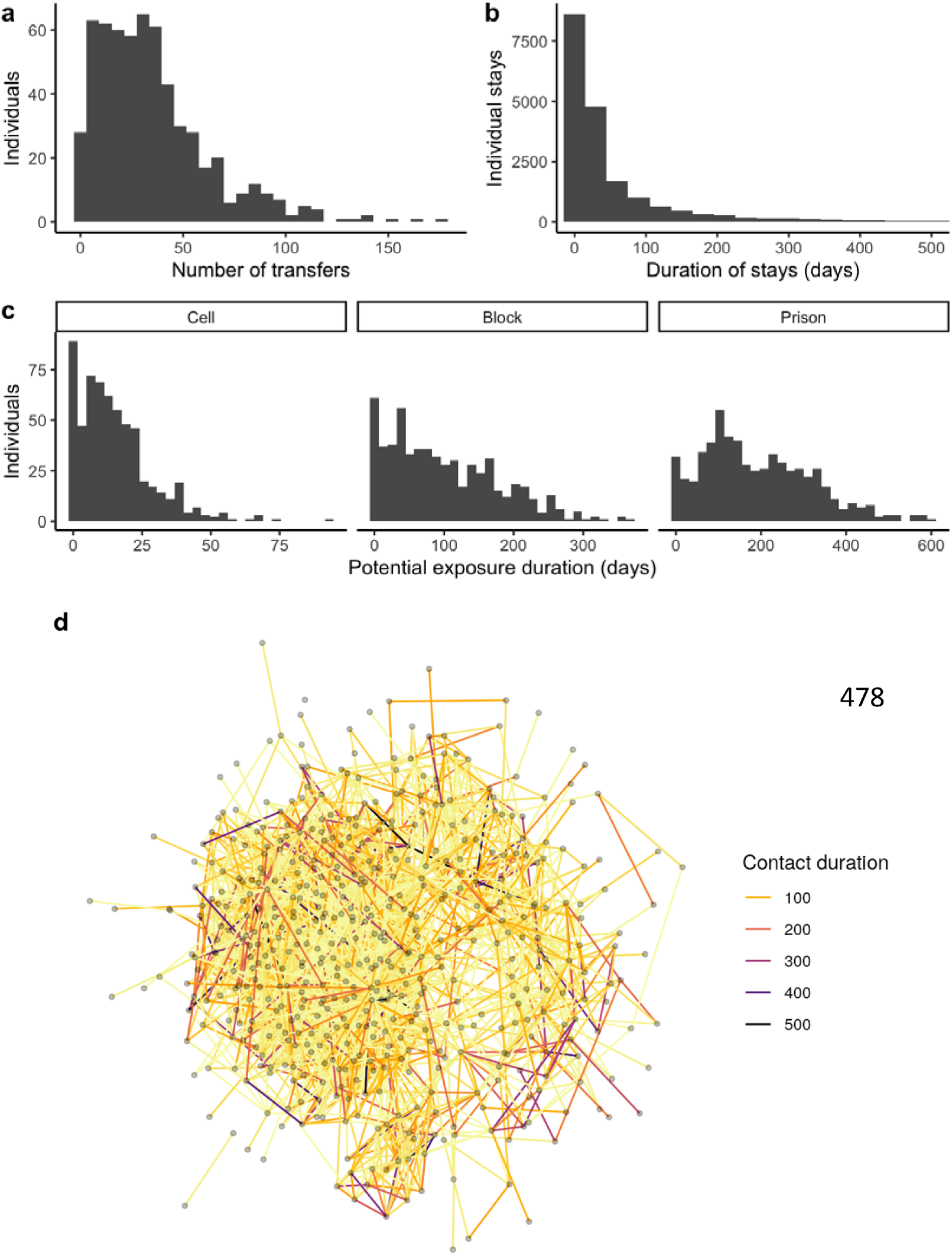
Frequent transfers in the Mato Grosso do Sul, Brazil state carceral system create highly connected networks. Histograms of a) the number movements recorded for each individual, including movements within prisons, between prisons, and to and from prisons, b) the duration of stay for each period of incarceration, and c) the duration of potential *M. tuberculosis* exposures, which we define as overlapping time and space prior to an individual’s TB notification, within the same cell, block of cells, or prison. d) Contact networks including all individuals with at least one cell-level potential exposure of one week or longer. Nodes indicate individuals, edges connect individuals with potential cell exposures of one week or longer and edge color indicates the duration of cell level exposure.

### Potential *M. tuberculosis* exposures are frequent and are associated with genomic cluster membership

To determine if known exposures within prisons predicted *M. tuberculosis* transmission linkages, we quantified potential exposures within cells, blocks of cells, and the prison as overlapping incarceration with another individual with notified TB prior to the recipient’s date of TB notification (Methods).

With that definition, 89% (528 of 595) of individuals with available incarceration data had at least one potential cell-level exposure, 93% (553 of 595) had at least one block-level exposure, and 97% (579 of 595) at least one prison-level exposure. Individuals had a median of 12 (IQR: 5-21) potential unique exposures to individuals of at least one week in cells, 76 (IQR: 32-138) in blocks, and 143 (IQR: 84-218) in prisons (Fig. 2).

For 561 of the 595 individuals with detailed incarceration histories, we sequenced high quality *M. tuberculosis* genomes from diagnostic cultures. Of these, 11 were classified as mixed strain infections, with more than one sub-lineage present, and we excluded these from subsequent analyses, as we were not able to reconstruct complete haplotypes. The majority (99.6%, 548/550) of single strain infections were in Lineage 4 and two isolates were from lineage 1 (sublineage 1.1.3.2) (Fig. 3). The most common sublineages were 4.1.2.1 and 4.4.1.1, including 172 and 121 isolates, respectively. To investigate potential transmission linkages more closely, we applied commonly used 5 and 12-SNP genetic distance thresholds. The majority, 66% (363 of 550) of individuals, fell into 32 clusters when applying a 5-SNP genetic distance threshold and 88% (484 of 550) of individuals fell into 37 clusters when applying a more liberal 12-SNP threshold (Supplementary Figure 1).

**Fig. 3.**
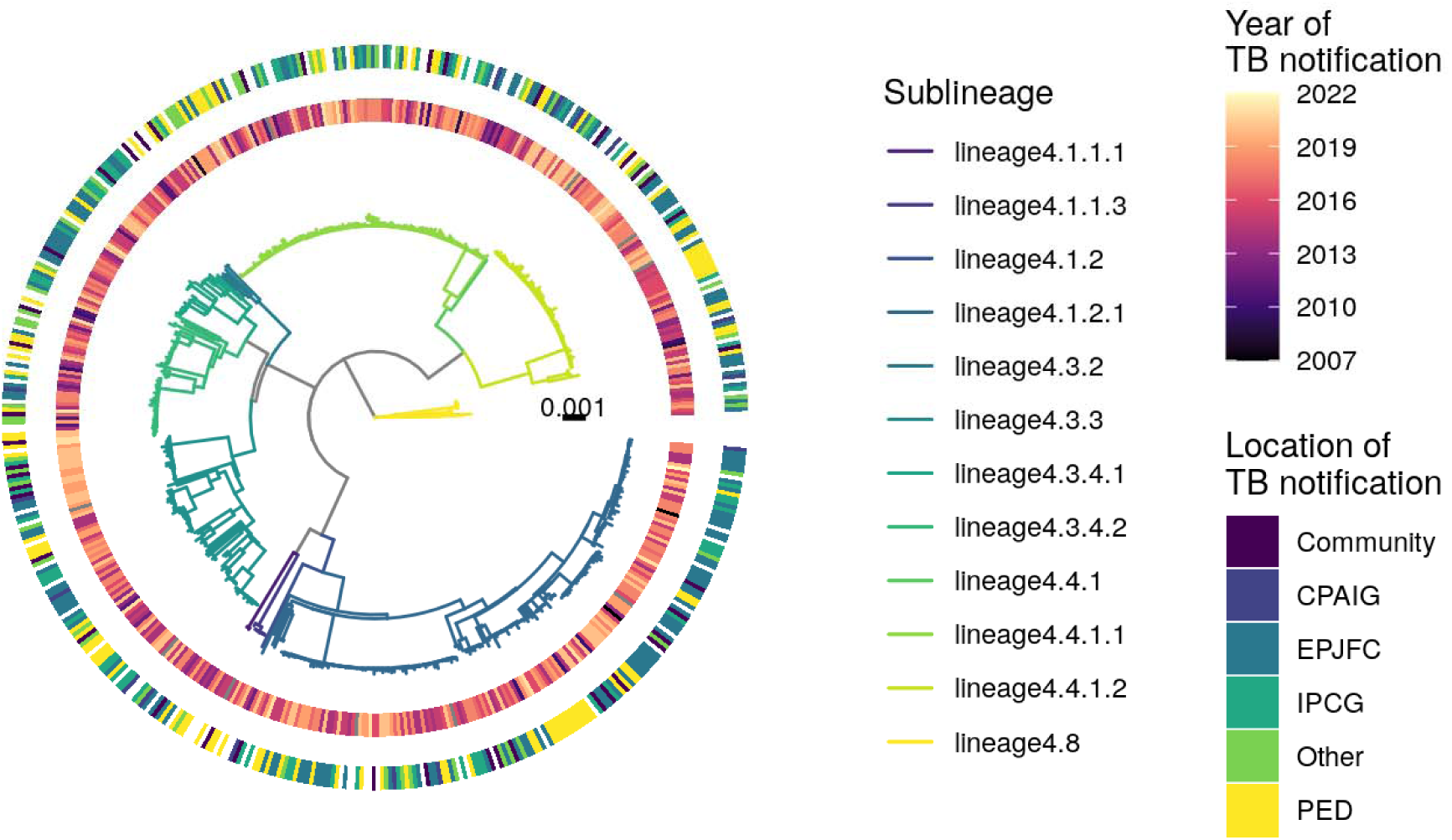
Maximum likelihood phylogeny of *M. tuberculosis* isolates infecting incarcerated individuals in Mato Grosso do Sul, Brazil. Phylogeny inferred with IQ-TREE and rooted on the 17 lineage 4.8 genomes from our study. The two *M. tuberculosis* lineage 1 genomes are excluded for visualization. Branch colors indicate sublineage and branch lengths represent substitutions per site. The inner ring indicates year of TB notification and outer ring indicates the location of TB notification. The largest prisons are specified; any prison with fewer than 15 notifications is labeled Other.

Of the 363 individuals infected with a clustered *M. tuberculosis* isolate, 329 (90.6%) had at least one cell-level potential exposure. Cluster membership (infection with an *M. tuberculosis* isolate falling in a genomic cluster defined by a 5-SNP threshold) was associated with at least one cell-level potential exposure (odds ratio (OR): 3.12, 95% CI: 1.39-7.33), in a logistic regression model.

### Cell, block, and prison exposures are associated with, but not predictive of *M. tuberculosis* **transmission linkage.**

To test if individual-level contacts within the prison were predictive of transmission linkages to other individuals, we fit a second logistic regression model for pairwise genomic cluster membership as a proxy for transmission, including the duration of potential pairwise exposures within cells, blocks, and prisons as predictors, and controlling for non-independence of paired data (Methods). Potential exposures were significantly associated with *M. tuberculosis* genomic clustering when using a 5-SNP threshold, resulting in an odds ratio of 1.07 (95% CI 1.04-1.10) for one month-long increase in duration of overlapping incarceration in a cell; OR: 1.01 (95% CI 1.007-1.02) for one month-long increase in days of overlapping incarceration in a block; and OR: 1.005 (95% CI 1.003-1.008), for month-long increase in days of overlapping incarceration in a prison (Fig. 4). The significant association between prison exposures and transmission remained under a more liberal, 12-SNP threshold for transmission linkage.

**Fig 4.**
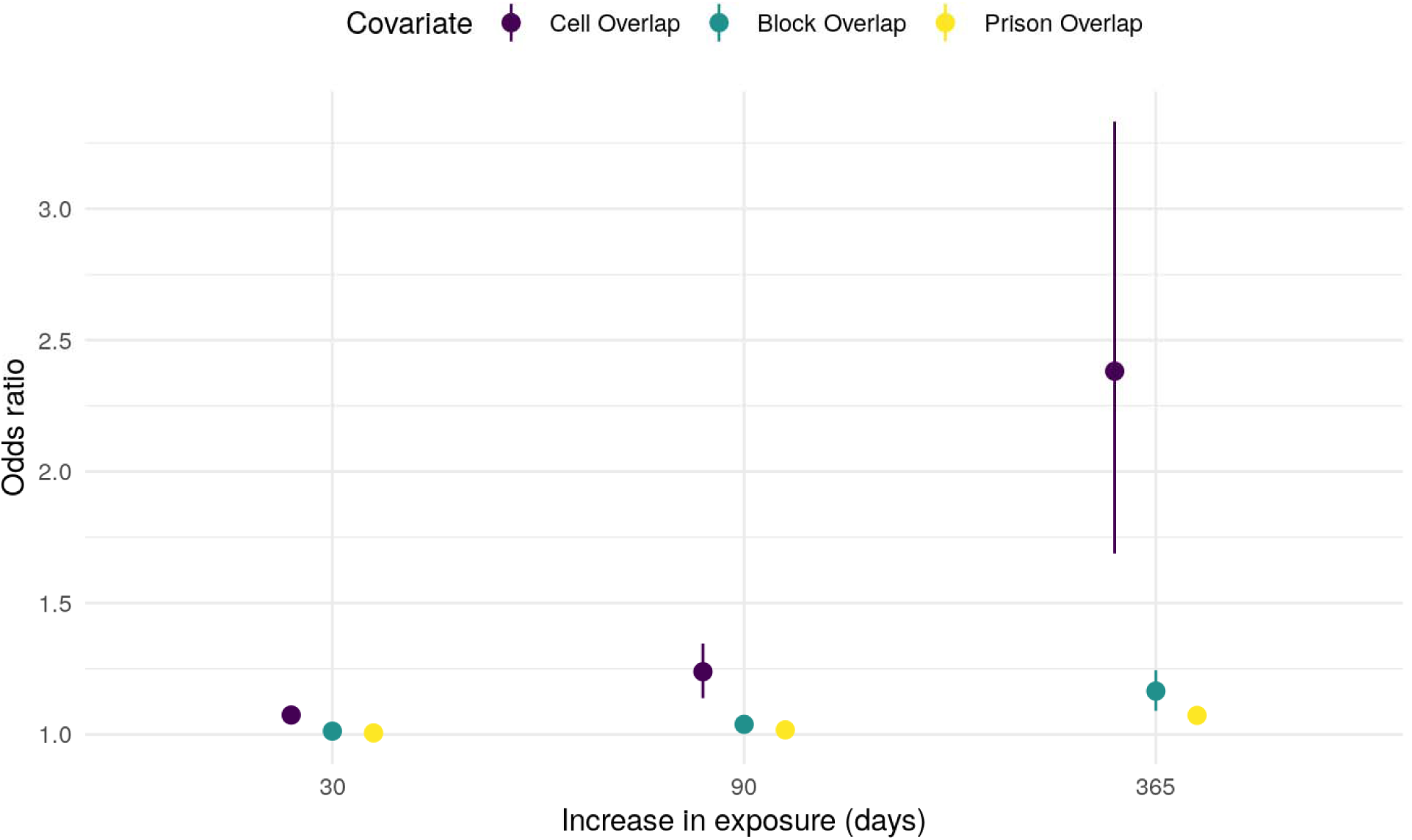
Odds ratio of genomic clustering as a function of prison exposures. The odds ratio for pairwise *M. tuberculosis* genomic clustering, with a 5-SNP threshold, as a function of days of exposure within a cell (purple), block (green), or prison (yellow), with whiskers indicating 95% confidence intervals.

However, exposure information did not perform well in discriminating between pairs of individuals linked in recent transmission. Predictions made by the above model including cell, block, and prison-level exposures had an area under the curve (AUC) of 0.51 for predicting membership in a genomic cluster defined by a 5-SNP genetic distance threshold (Supplementary Methods).

### Multiple *M. tuberculosis* clones co-circulate within prisons

We hypothesized that the poor predictive value of prison exposures for genomic linkage might reflect the fact that in such high-incidence congregate settings, individuals might have multiple exposures to *M. tuberculosis*, with the effect that any single exposure might not predict transmission linkage. Among the 472 individuals at least one week-long potential cell exposure, 315 (66.7%) had at least one potential exposure to an individual within the same genomic cluster; 229 (48.5%) had more than one potential exposure to an individual within the same genomic cluster, 285 (60.4%) had exposures to both individuals within and outside the genomic cluster, and 157 (33.3%) had exposures only to individuals within a different genomic cluster, when applying a 5-SNP threshold for clustering. Of the 363 individuals infected with a clustered isolate, 153 people did not have any cell exposure with other individuals in the same 5-SNP genomic cluster.

Additionally, in the three prisons in which our group conducted systematic surveillance, and which have the highest genomic coverage, we found evidence of co-circulation of multiple sublineages over time (Fig. 5). We observed up to 8 sublineages co-circulating within a single year in Estabelecimento Penal “Jair Ferreira de Carvalho” (EPJFC), a closed prison in the state capital, Campo Grande; and 7 in both (Instituto Penal de Campo Grande (IPCG), a closed prison in the state capital, and Penitenciária Estadual de Dourados (PED), a closed prison in the city of Dourados.

**Fig 5.**
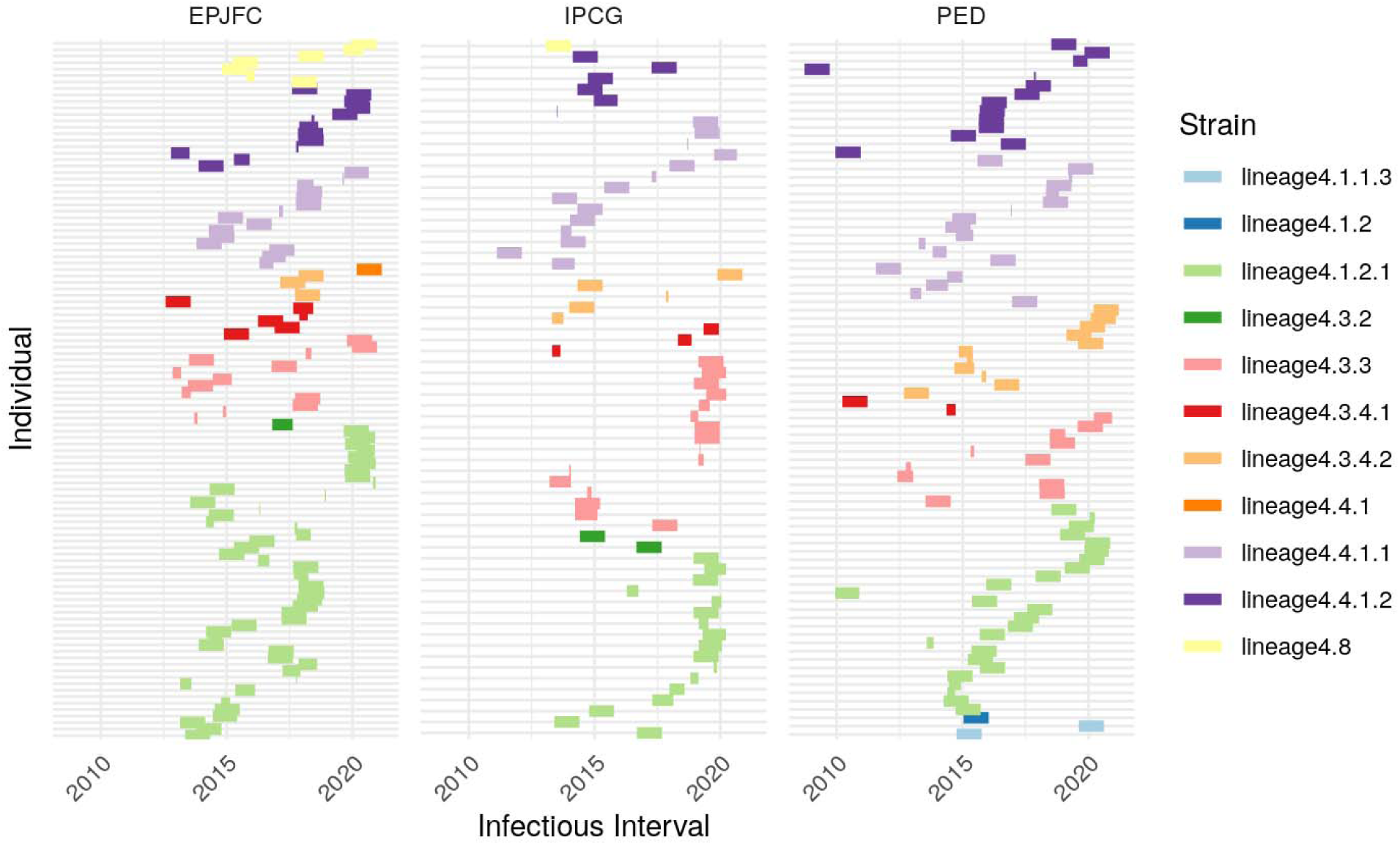
Multiple *M. tuberculosis* clones co-circulate in high-incidence prisons. Lines indicate individuals infected with TB and panels indicate location of incarceration. Line color indicates *M. tuberculosis* sub-lineage. Lines are shaded during a potential infectious period, one year prior to TB notification. The three prisons in which active TB screening was conducted are included here.

### Contact heterogeneity within genomic clusters

In the largest genomic clusters, including 102 and 50 sequences, 73% and 52% of individuals had at least one reported cell exposure to another individual within the same genomic cluster, respectively (Fig. 6). We observed contact heterogeneity, with the 10% most connected individuals responsible for 33% of contact time and 27% of unique people contacted in the largest cluster and 40% of contact time and 29% of unique people contacted in the second largest cluster (Fig. 6).

**Fig 6.**
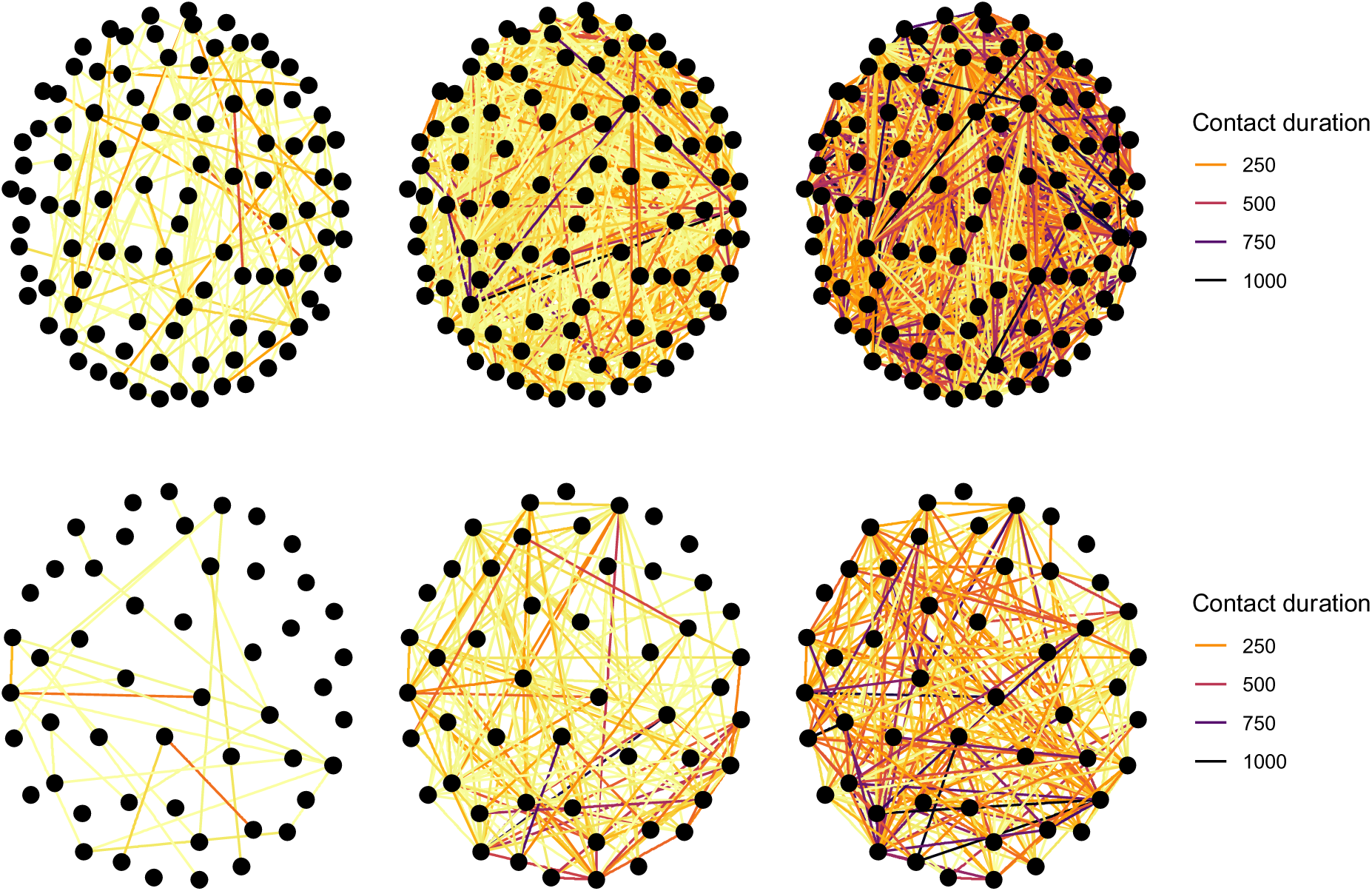
Contact networks for the two largest genomic clusters. Nodes indicate individuals and edges connect individuals with potential exposures within cells (left column), blocks (center column), and prisons (right column). Edge color indicates duration of exposure in days. The top row indicates the contact network for the largest genomic cluster (102 sequences) and bottom row indicates the second largest genomic cluster (50 sequences).

## Discussion

We found that both close contacts (incarceration in the same prison cell) and more distant contacts (incarceration within the same prison), are significantly associated with pairwise genomic clustering, a proxy for transmission, in a state prison system with extremely high incidence of TB. The frequent transfer of incarcerated individuals within and between prisons generates a significant number of potentially infectious contacts and potential exposures to several co-circulating *M. tuberculosis* genotypes. The result of the high force of infection and frequent transfers is that contact tracing investigations may not be an effective tool in identifying sources of transmission.

Previous studies have also employed molecular methods to identify sources of recent *M. tuberculosis* transmission. Studies in several settings have found that only approximately 10-30% of cases can be attributed to transmission from a known household contact and have concluded that transmission is largely driven by community transmission^2–7^. In high transmission settings, such as KwaZulu Natal, South Africa, even household contacts are not genetically linked^3^. While fewer studies have focused on transmission within carceral settings, we similarly find that known close contacts (here, cell-level contacts) alone could not explain most transmission linkages. In this setting, exposures of any type are associated with increased odds of genomic clustering, suggesting that both close and distant contacts play a role in driving transmission.

Our results highlight that the frequent transfer of people within prisons and between prisons creates large contact networks in which almost all individuals have many potentially infectious contacts. This contact network is further expanded by the many prison workers and visitors who pass through prisons every day and whose movements are not included in the carceral movement database. The frequent movement of individuals by the prison system is compounded by severe overcrowding of prisons and individual cells. Nationally, Brazil’s 1,384 prisons are at a 174% occupancy rate^9^. In one of the prisons included in this study, up to 69 people were incarcerated per cell, a total of 0.3 m^2^ area per person. Most prison cells fail the World Health Organizations standards for the minimum airflow required to prevent airborne infections ^12^.

The impact of the high force of infection and close contact network is that (1) that recorded cell-level exposures do not explain all transmission, as many individuals infected with a clustered *M. tuberculosis* isolate do not have known cell-level contacts with potential donors infected with an isolate from the same cluster; and (2) this may, in part, reflect contact saturation, in which repeated contacts with previously infected individuals do not lead to transmission and are “wasted.” ^25^ In a previous modeling study, contact saturation, in conjunction with superspreading, explained the low proportion of transmission attributable to household contacts despite their relatively high proportion of actual contact time^25^.

Our findings that both close and more distant contacts contribute to the odds of *M. tuberculosis* genomic cluster membership and transmission, support the use of agnostic interventions to suppress uncontrolled transmission across the entire prison population. Previous studies have demonstrated that systematic screening in prisons can improve TB detection^24^ and modeling studies show that TB screening at prison entry, exit, and for the entire population at least annually can lead to early diagnosis and reduce TB incidence, both within prisons and in surrounding communities^26,27^. TB preventive treatment (TPT) for individuals at high risk of disease is one of the key recommendations of the World Health Organization’s End TB Strategy^28^. Studies have demonstrated that short-course TPT is widely accepted and has high completion rates in prisons and jails^14^. However, the most recent WHO recommendations for priority use of TPT do not include incarcerated populations at high risk of infection, where TPT use is only a “conditional recommendation.”^29^ Evidence also indicates that structural interventions including reducing severe overcrowding, improving ventilation, expanding time outdoors, and improving access to nutritious food and routine medical care can reduce infection risk^12^. In high incidence, highly connected settings, directing interventions towards known close contacts alone may not sufficiently interrupt transmission linkages. Additionally, contact tracing is resource-intensive and challenging in congregate settings and particularly in carceral systems, where access to movement data is limited and not routinely available to public health.

The most direct way to reduce the infectious disease risk created by incarceration is to reduce the number of people sent to high incidence settings, by reducing unprecedented incarceration rates and decreasing reliance on incarceration. A dynamic transmission model calibrated to incarceration and TB data from several Latin American countries found that reductions in prison admissions and the duration of incarceration would reduce TB incidence by more than 10% in countries including Brazil^30^.

Our study has several limitations. First, the incarceration database was only accessible through manual query, and we could only collect incarceration histories for individuals with TB. We therefore do not measure TB transmission risk, but instead odds of genomic linkage among individuals who do have TB. Second, we underestimate ongoing transmission because of incomplete genomic sampling of all infected individuals, including those who are asymptomatic or do not produce sputum, potentially leading us to underestimate transmission risk. Third, there is some missing information due to reporting inconsistencies in the prison location data. Our team extracted incarceration histories manually and resolved ambiguities in location names and we do not expect that this would bias our results. Fourth, we followed a standard approach in *M. tuberculosis* genomic epidemiology: *M. tuberculosis* isolates were cultured and sequenced with short reads to generate single consensus sequences for each individual infection. Because of the limitations of short read sequence data, we were not able to reconstruct multiple genomes from individuals identified as multiply infected who may be contributing to onwards transmission. The relatively low prevalence of mixed infections in this study reflects our initial criteria for selecting individuals diagnosed with TB and with high-quality *M. tuberculosis* genomes for study inclusion and our use of solid cultures. Finally, we study a specific carceral setting in Central West Brazil. While evidence from other congregate settings suggest similarly high force of infections, our findings may not be entirely generalizable.

Here, we characterize patterns of *M. tuberculosis* transmission in a criminal justice system with extremely high incidence of TB. Close exposures within prison cells were associated with pairwise *M. tuberculosis* genomic clustering, a proxy for transmission. We also found that the frequent transfer of individuals results in individuals having many potential exposures. Together, this suggests that contact tracing investigations may not be useful in this setting. Our findings highlight the urgent need to reduce the force of infection within prison systems to protect the health of extremely vulnerable populations.

## Data Availability

Sequence data for samples meeting quality filters are available on the Sequence Read Archive (SRA), in BioProject PRJNA671770.

https://www.ncbi.nlm.nih.gov/sra

## Notes

### Competing Interest Statement

The authors have declared no competing interest.

