## Supplementary Information for "*M. tuberculosis* transmission dynamics in congregate settings: a genomic epidemiology study"

**Supplementary Methods**

**Variant identification**

We previously conducted a variant identification experiment to compare commonly used mapping and variant calling algorithms in *M. tuberculosis* genomic epidemiology^31^ and found that the combination of the *bwa*^32^ mapping algorithm and *GATK*^33,34^ variant caller routinely minimizes false positive variant calls with minimal cost to sensitivity as compared to other tool combinations^35^, in particular, when the PE/PPE genes are excluded. We therefore used this combination of tools in our pipeline.

Briefly, we trimmed low-quality bases (Phred-scaled base quality < 20) and removed adapters with Trim Galore v. 0.6.5 (stringency=3)^36^. We used CutAdapt v.4.2 to further filter reads (--nextseq-trim=20 --minimum-length=20 --pair-filter=any)^37^.To exclude potential contamination, we used Kraken2 to taxonomically classify reads and removed reads that were not assigned to the *Mycobacterium* genus or that were assigned to a *Mycobacterium* species other than *M. tuberculosis*^38^. We mapped reads with *bwa* v. 0.7.15 (*bwa* mem)^32^ to the H37Rv reference genome (NCBI Accession: NC_000962.3 [https://www.ncbi.nlm.nih.gov/nuccore/NC_000962.3]) and removed duplicates with *sambamba*^39^. We called variants with GATK 4.1 HaplotypeCaller^33^, setting sample ploidy to 1, and GenotypeGVCFs, including non-variant sites in output VCF files. We included variant sites with a minimum depth of 10X and a minimum variant quality score 40 and constructed consensus sequences with bcftools consensus^40^, excluding indels. We excluded SNPs in previously defined repetitive regions (PPE and PE-PGRS genes, phages, insertion sequences and repeats longer than 50 bp)^41^. We identified sub-lineage and evidence of mixed infection with *TBProfiler v.4.2.0*^42,43^, which is based on the identification of >1,000 [lineage-specific SNPs](https://github.com/jodyphelan/tbdb/blob/master/barcode.bed).

**Phylogenetic analysis**

We constructed full-length consensus FASTA sequences from VCF files, setting missing genotypes to missing, and used SNP-sites to extract a multiple alignment of internal variant sites only^44^. We used the R package *ape* to measure pairwise differences between samples (pairwise.deletion=TRUE)^45^. We selected a best fit substitution model with ModelFinder^46^, implemented in IQ-TREE multicore version 2.2.0^47^, and fit a maximum likelihood tree to the alignment of SNP sites with IQ-TREE, with 1000 ultrafast bootstrap replicates, with an ascertainment bias correction for using only variant sites ^47,48^.

**Statistical analysis**

We assessed logistic model performance by comparing model predictions with observed data with a receiver operating characteristic curve, summarized by area under the curve (AUC) with the R package *yardstick^23^.* In addition to the model described in the main text, we fit a model that with an additional term, an indicator variable for any prison cell exposure, reasoning that this might account for possible zero-inflation in our data, as most pairs of individuals have 0 days of overlapping exposure. In this model, the covariate for the cell exposure indicator variable was not significantly different from zero.

To quantify the number of co-circulating *M. tuberculosis* sublineages within each prison, we considered an incubation period (time from infection to TB diagnosis) of one year and counted the unique sublineages represented each year (Fig. 5).

**Supplementary Figures**

**Figure S1. Distribution of genomic cluster sizes.** Histogram of the number of *M. tuberculosis* sequences in each genomic cluster (including 2 or more genomes) identified after applying alternative 5- and 12-SNP distance thresholds.

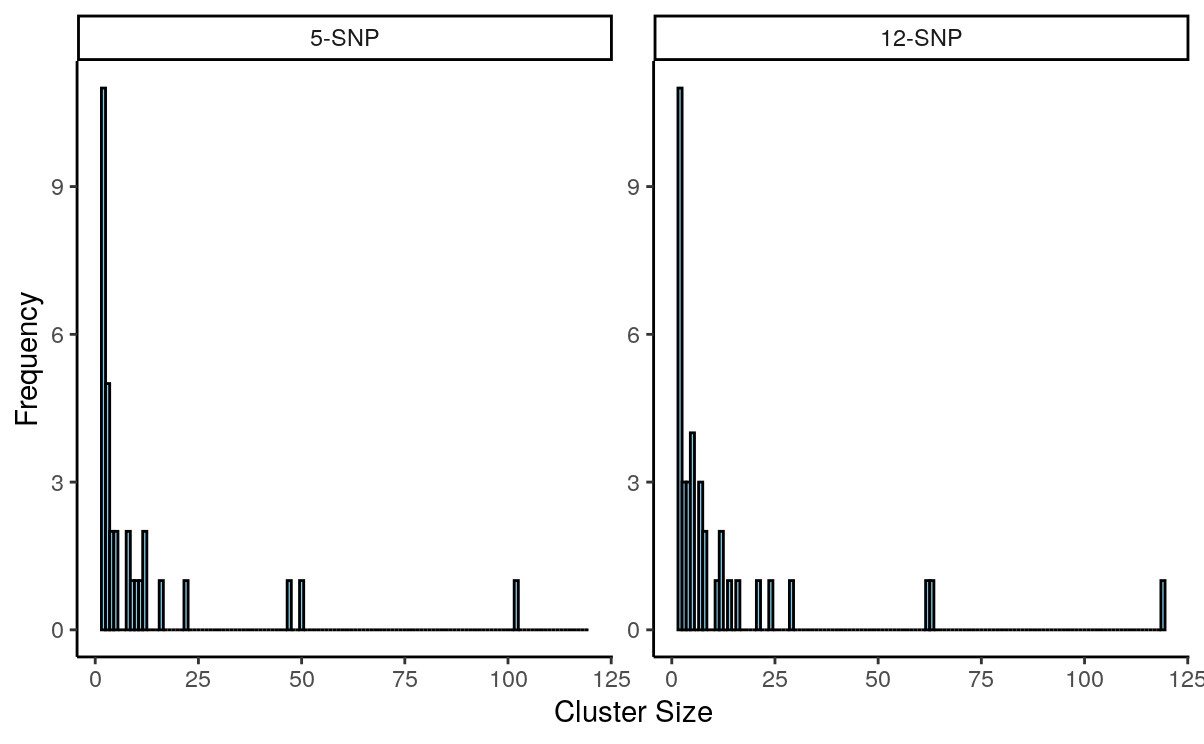
